# A modelling approach for the analysis of decisions in Home Health Care at multiple planning levels

**DOI:** 10.1101/2025.11.06.25339521

**Authors:** Luca Grieco, Martin Utley, Sonya Crowe

**Affiliations:** Clinical Operational Research Unit, Department of Mathematics, University College London, London, United Kingdom

**Keywords:** Home health care, OR in health services, Optimisation, Strategic planning, Decision making, Synthetic data

## Abstract

The sustainable delivery of Home Health Care (HHC) is increasingly important as ageing populations and rising demands place pressure on health and social care systems. HHC involves complex decision-making across strategic, tactical, and operational levels, often constrained by limited resources such as workforce availability.

While Operational Research (OR) has been widely applied to support operational decisions in HHC, recent reviews highlight a lack of focus on strategic and tactical planning, and limited recognition of the hierarchical structure linking decisions across levels. This undermines the potential of OR to inform real-world planning effectively.

To address these gaps, we propose a modelling approach enabling consistent analysis of decisions across planning levels. We defined a modular approach for analysing hierarchies of decisions and accounting for cascade effects. Then, we applied those principles by developing a configurable tool in R, comprising a synthetic data generator and a suite of optimisation and heuristic routines.

We illustrate the benefits of this approach through a case study. Our results demonstrate the value of this structured modelling approach for informing decisions in HHC. The emphasis we gave on modularity facilitated the development of an analysis tool that can be easily adapted to different hierarchies of decisions and settings.

## 1. Introduction

Efficient deployment of resources to deliver home-based care is critical for the sustainability of health care systems, given the trend towards ageing populations with more complex needs and the increasing pressures that health and social care services are experiencing (Kingston et al. 2018, Stevenson and Mutebi 2024).

Service delivery in Home Health Care (HHC) is a complex process, with local authorities, commissioners, provider organisations and health care workers involved in making decisions at strategic, tactical and operational levels (Hulshof et al. 2021). Challenges in such decisions are further exacerbated by the scarcity of key resources, including the workforce, and sustained increase in demand (NHS England 2023).

Operational Research (OR) approaches have been widely used to model operations and support decision-making in HHC. However, recent literature reviews of OR applied to HHC reported an abundance of work focusing on operational decisions and less work at strategic and tactical levels, and little explicit recognition of their hierarchical nature (Gutiérrez and Vidal 2013, Sahin and Matta 2015, Grieco et al. 2021), which undermines the potential of OR approaches to support real-life decisions. More recently, Chabouh et al. 2023 surveyed the literature with a specific focus on strategic and tactical decisions, reporting some increase in the number of publications covering decisions such as network design and resource dimensioning, but still an overall lack of studies for longer-term planning and particularly of such studies that also consider uncertainty in service delivery. They also observed how the diversity of contexts characterising HHC systems makes comparisons of different modelling approaches particularly challenging, and that availability of benchmark instances would make comparative analyses of models and methods easier.

In this study, we addressed these research gaps by developing a modelling approach that can be used to analyse decisions in HHC consistently across planning levels. The contribution of this paper is twofold: firstly, we define an approach to structuring the analysis of sequential decisions at strategic, tactical and operational levels in HHC, therefore accounting for hierarchical structures and cascade effects; secondly, as an example implementation of our approach, we provide a configurable tool (developed in R programming language) consisting of a synthetic data generator and a suite of interlinked optimisation/heuristic routines, and illustrate its use through a case study inspired by a question faced by a collaborating HHC organisation in London, UK.

In what follows, we firstly give an update to Grieco et al. 2021’s literature review of OR applied to HHC (section 2), and next present our modelling approach (section 3). We then describe the instantiation of our approach around a question of interest to our reference HHC organisation (section 4): we present the decision framework of interest, the generation of realistic synthetic data for use in our analysis pipeline, the methods for the solution of the corresponding decision problems, and the parameterisation of our models for the analysis of the case study. We conclude with a discussion about the advantages and limitations/challenges of our modelling approach, and possible future directions (section 5).

## 2. Literature review

To assess whether any of the gaps identified by Grieco et al. 2021 have been addressed in the last few years, we used the same search strategy used in that literature review to identify articles on the application of OR in HHC published since September 2018 (date of the search run by Grieco et al. 2021). The papers were screened by title and abstract using the same inclusion and exclusion criteria described in Grieco et al. 2021, and then classified based on their main contribution in an attempt to identify publications that would address decision making in HHC at multiple planning levels.

Of the 476 articles identified, we discarded 9 literature reviews and 201 out-of-scope publications. The vast majority (about 90%) of the remaining studies exclusively focused on decisions at operational levels, while 19 papers addressed strategic or tactical decisions.

The remaining 11 studies addressed decision making at multiple planning levels (see Table 1 for brief summaries of these studies). A recurring topic among these papers was the design of HHC networks (i.e. deciding which HHC centres or pharmacies to open) while optimising a combination of operational decisions (rostering/allocation/scheduling/routing). All of them propose optimisation models that address decisions at different planning levels simultaneously, usually using a multi-stage optimisation approach, and develop algorithms to cope with the computational complexity of the problem.

**Table 1.**
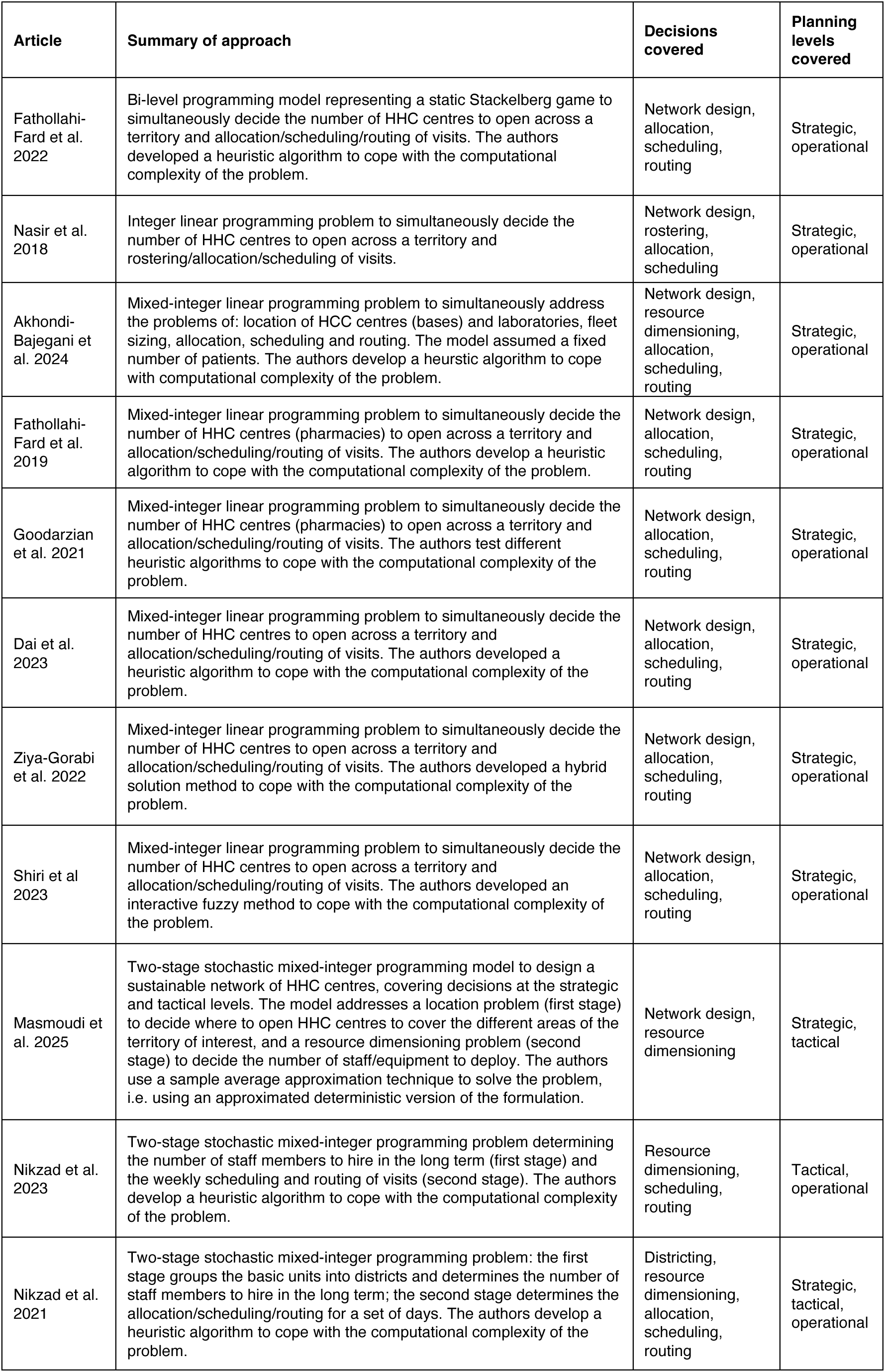
Recent studies (published since September 2018) addressing decision making in HHC at multiple planning levels.

These findings confirm the trends identified by the literature reviews mentioned in the previous section. In particular, we did not find any additional studies exploiting the hierarchical structure of decisions in HHC in their analyses.

## 3. Modelling approach

Our modelling approach consists of characterising a set of decision problems to be solved in sequence according to a given hierarchical ordering, with the output of one decision problem feeding into subsequent decision problems, and analysing them by exploiting an underlying dataset that accounts for the characteristics of the health economy of interest.

We organise the decisions of interest into a modular structure. Each decision belongs to a *module* in the hierarchy (Figure 1a). Modules are interconnected in logical sequences depending on how decisions affect each other (Figure 1b). A module could also be formed of a set of decisions, as long as they are made simultaneously (e.g. visit scheduling and staff-to-visit allocation). As a result, we suggest analysis frameworks in which decisions that happen at different planning levels always belong to different modules in a hierarchy.

**Figure 1.**
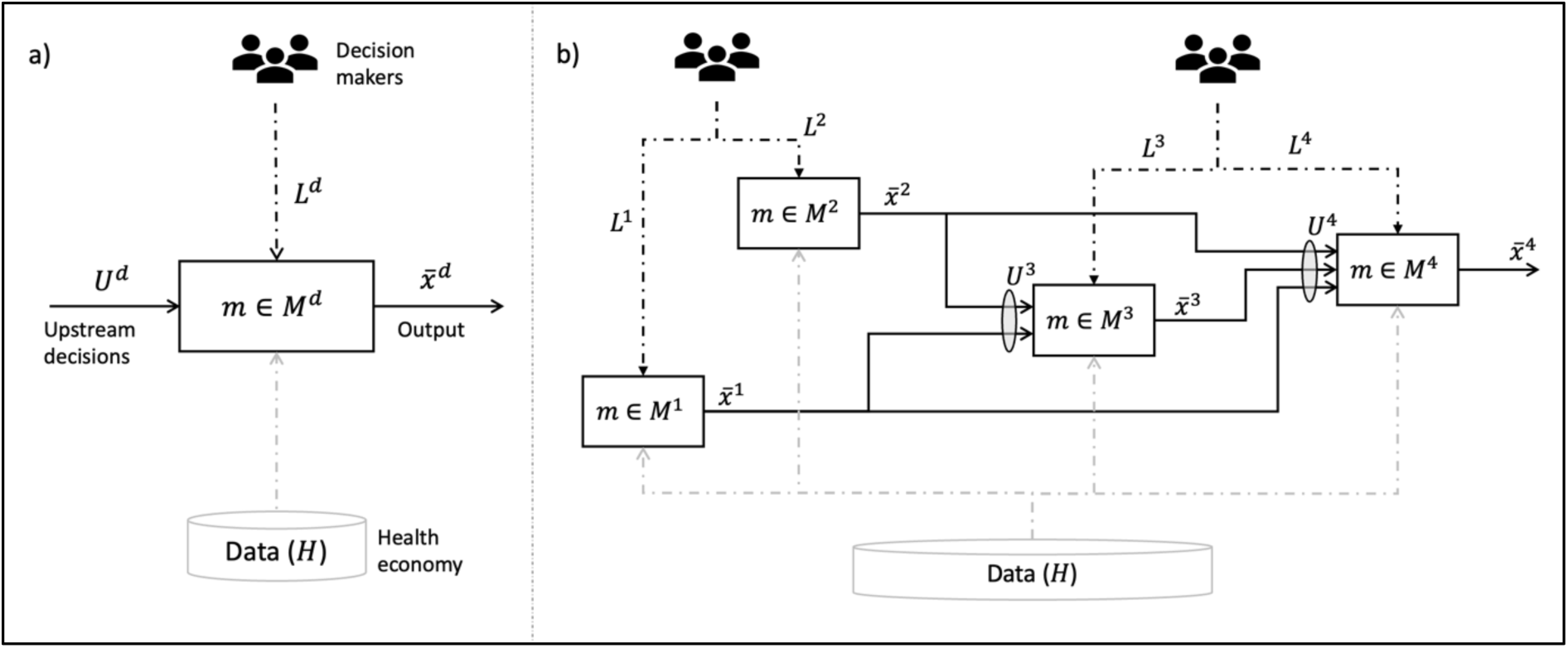
A generic module in our modelling approach (a) and an example of a decision diagram formed of four decisions (b).

The formalisation that we provide here can serve as a guide for specific implementations of case studies, such as the one described in the next section.

Let the collection 𝒟 represent our decisions of interest, where each 𝑑 ∈ 𝒟 can be either a single decision or a set of decisions to be made simultaneously. For each 𝑑, we need to find values for a vector of decision variables 𝑥̅^d^ ∈ ℝ^n^. There might be different ways to determine 𝑥̅^d^, for instance using an optimisation method, a heuristic algorithm, or a set of instructions mimicking the procedure currently adopted by the decision maker. We can therefore define a set of methods 𝑀^d^. Different sets of analyses of the same hierarchy of decisions might use a different method 𝑚 ∈ 𝑀^d^ to produce the same type of output 𝑥̅^d^ ∈ ℝ^n^, therefore enabling consistent comparisons between different decision-making approaches.

A key aspect for a systematic and reliable analysis of the hierarchy of decisions is the quality of the flow of information between the models. Consistency in this flow can be achieved by using a unique and self-contained dataset representing the health economy from which parameters can be estimated for all the decision problems of interest. We denote all the data related to the health economy with 𝐻. These translate into fixed parameters in each of the decision problems of interest. For instance, if part of 𝐻, sampled locations of patient homes can be used for estimating distances of wards in deciding the composition of the different territorial units (districts) at strategic level, as well as to directly parameterise a routing algorithm at operational level.

It is also important to keep the representation of the health economy separate from other features of the decision-making process, such as arbitrary assumptions by decision makers and exogenous elements that result from previous decisions in the analysis sequence.

Therefore, we can define:

- A set of “levers” 𝐿^d^ that decision makers can act upon in making the set of decisions 𝑑. These could be parameters that can affect or scope a decision and that do not depend on upstream decisions or on underlying data (e.g. minimum proportion of satisfied demand to ensure, what goal to optimise, etc.).
- Implications from upstream decisions in the hierarchy. We denote these as 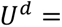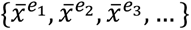, where 𝑒_1_, 𝑒_2_, 𝑒_3_, … ∈ 𝒟 are decisions happening upstream of 𝑑, and they correspond to the vectors of decision variables for those decisions that are used as input for 𝑑, with values established at previous steps in the cascade.

Finally, we measure the performance associated with 𝑥̅^d^ through a 𝑘-dimensional vector 𝛱^d^(𝑥̅^d^) ∈ ℝ^k^ that we can use to compare the different scenarios analysed.

In the next section, we will show how we put the above principles in practice through a case study inspired by a question faced by a collaborating home health care organisation.

## 4. Case study

Our case study is based on a question faced by a health care organisation providing home health care services in a borough of London. We have simplified and anonymised the features of the health economy of interest for the sake of presentation.

### 4.1 Settings

#### 4.1.1 London borough and question of interest

We consider the territory shown in Figure 2, split into ten administrative wards (basic units), covering an area of 15 square kilometres and with an expected number of 500 patients to receive HHC visits over a planning horizon of 20 weeks. This corresponds to a subset of (anonymised) wards belonging to our reference borough.

**Figure 2.**
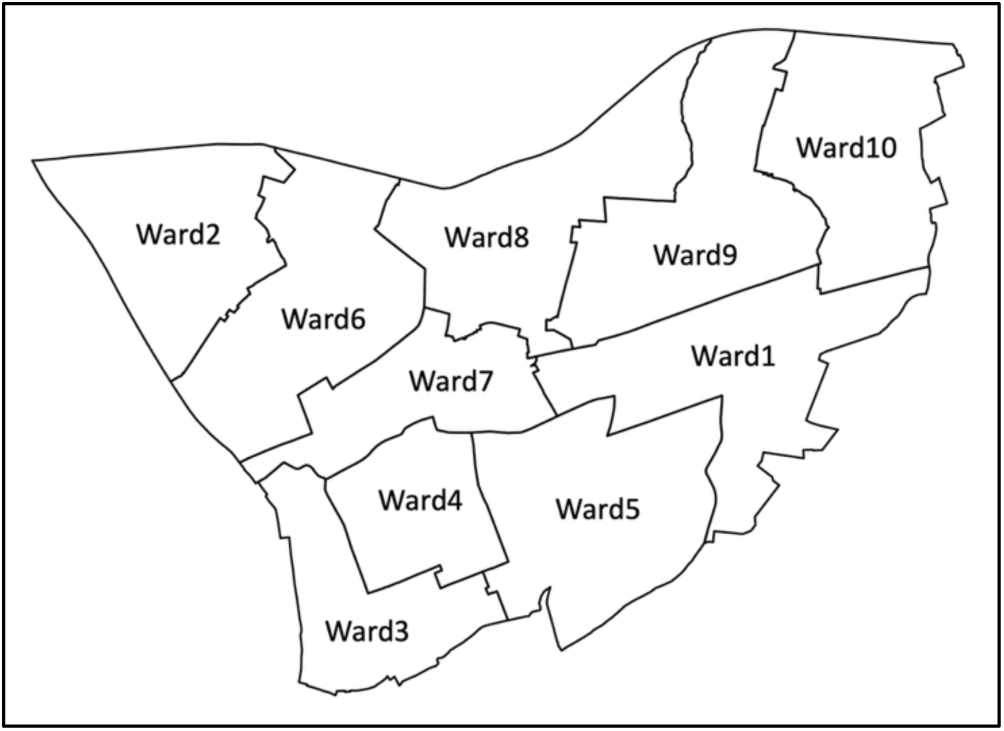
Territory of interest, with subdivision into ten basic units.

In our case study, the reference organisation provides home visits for two types of services: District Nursing (DN) and Physiotherapy (PT). Depending on their role, health care workers can be assigned to visits involving subsets of the activities listed in Table 2.

**Table 2.**
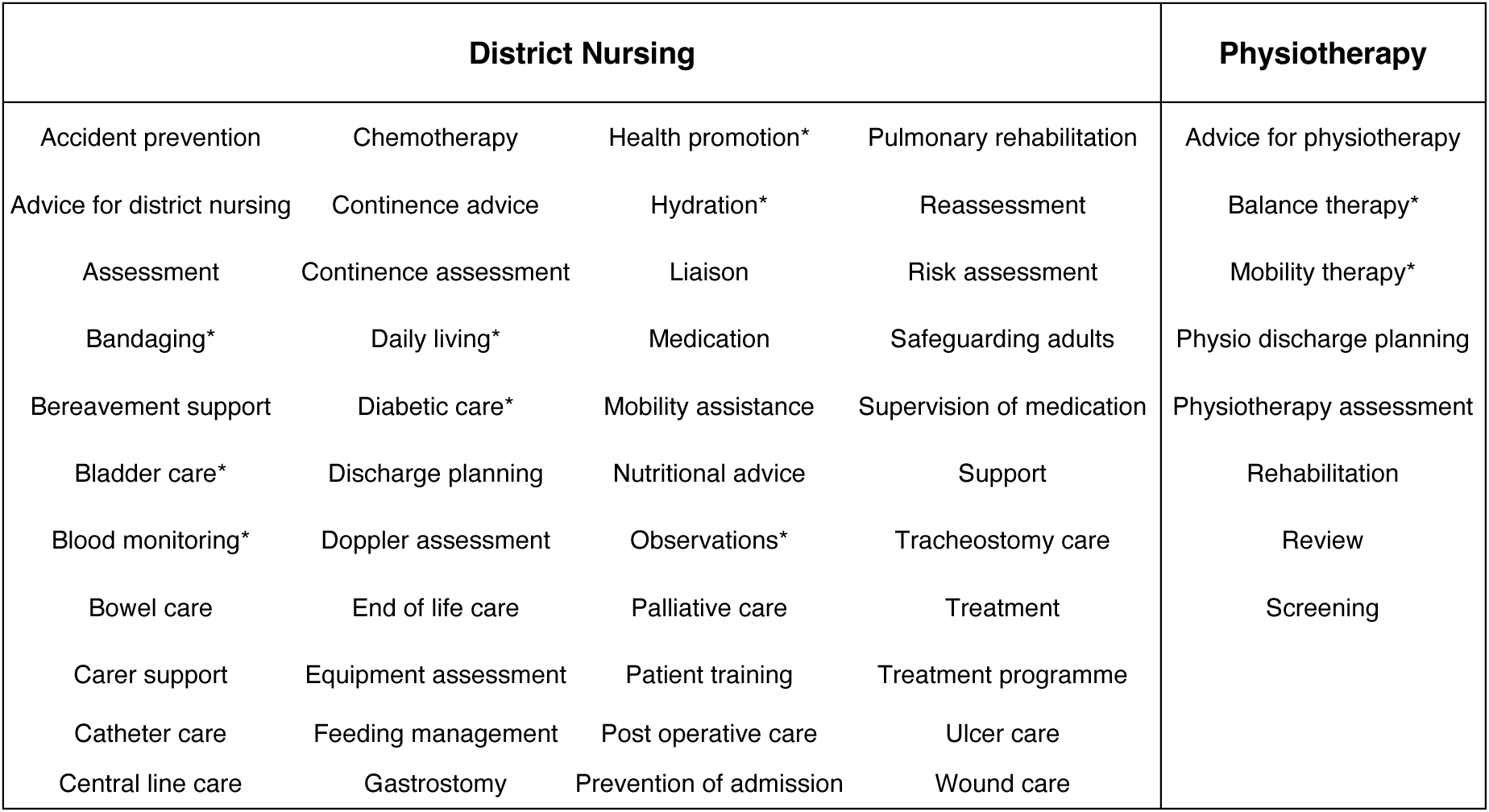
List of home-based activities considered in this case study. Asterisks indicate activities that could be conducted by workers in the new Hybrid Assistant role assessed in our analysis.

For this case study we considered a situation where a decision maker wants to assess whether to introduce a new workforce role (Hybrid Assistant) in the organisation. We assume that the organisation currently provides the two services separately: district nurses only undertake DN activities and physiotherapists only undertake PT activities. The district nurse and the physiotherapist roles are characterised by different seniority levels. We consider the three levels {“Junior”, “Mid-level”, “Senior”}, with varying hourly salary rates as summarised in Table 3. The new Hybrid Assistant role would be allowed to conduct a subset of activities (cf. Table 2) that would normally be conducted by either junior district nurses or junior physiotherapists, therefore visits conducted by workers in this hybrid role might involve a combination of DN and PT activities. Supplementary File 1 provides detailed lists of which activities can be conducted by workers in each role.

**Table 3.** List of roles considered in our case study. The Role ID column reports the identifiers we used in our case study results.

| Role category | Seniority level | Hourly salary | Role ID |
| --- | --- | --- | --- |
| District Nurse | Junior | £10 | DN_L |
|  | Mid-level | £14 | DN_M |
|  | Senior | £21 | DN_H |
| Physiotherapist | Junior | £10 | PT_L |
|  | Mid-level | £14 | PT_M |
|  | Senior | £21 | PT_H |
| Hybrid Assistant | Junior | £12 | HA_L |

Informed by discussions with our collaborating HHC organisation, the motivating hypothesis for this analysis is that, with activities allowed to be conducted by junior staff being the most common activities in HHC, introducing the Hybrid Assistant would enable a decrease in the number of visits needed to cover the demand, potentially translating into decreased travel costs as well as decreased workloads due to a more efficient packing of activities into visits. This might also improve patient experience that could potentially be characterised by a smaller number of distinct visits and a higher level of continuity of care.

#### 4.1.2 Decision hierarchy of interest

In the context of our collaborating HHC organisation, introducing the hybrid role is a strategic level decision involving the definition of workforce roles and HHC packages, but its effects in terms of efficiency and patient experience are also measurable at the point of service delivery (operational level). Therefore, in this analysis we showcase how our modelling approach could be exploited to inform strategic decisions based on their envisaged effects on downstream decisions in the hierarchical structure depicted in Figure 3.

**Figure 3.**
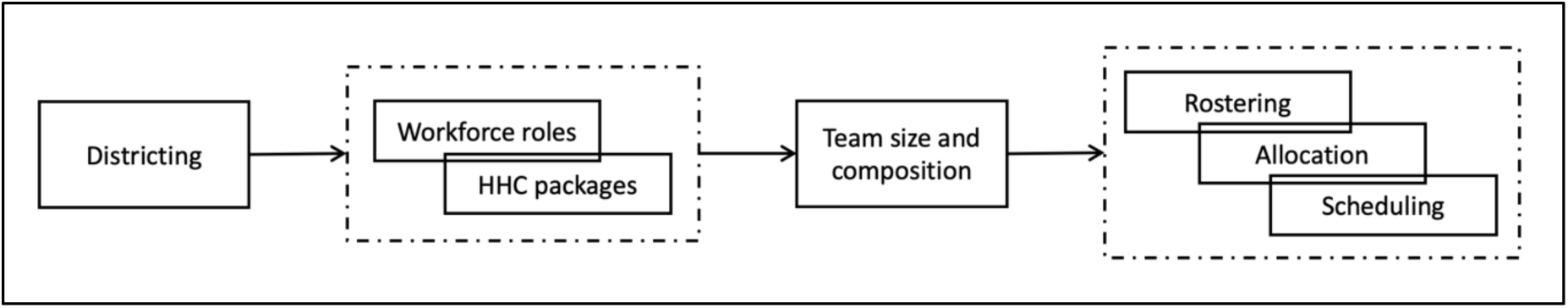
Decision hierarchy analysed in our case study, based on the decision framework proposed by Grieco et al. 2021. The decisions “Districting”, “Workforce roles” and “HHC packages” are at a strategic level and they influence decisions at a tactical level (“Team size and composition”) and at an operational level (“Rostering”, *“*Allocation*”,* Scheduling”). Dashed boxes denote sets of decisions to be made simultaneously, therefore belonging to the same module.

Table 4 provides a definition of each decision problem in the hierarchy. According to our modelling approach, for each module in the hierarchy of decisions, we defined a method to obtain a solution which we describe in section 4.3.

**Table 4.** Definition of the decision problems of interest used in our analyses (cf. Grieco et al. 2021).

| Decision problem | Definition |
| --- | --- |
| Districting | Given a set of basic units (i.e. wards) defining the territory (i.e. borough) of interest, group those wards into a pre-defined number of clusters, each representing a district |
| Workforce roles & HHC packages | Define visit types as combinations of DN and/or PT activities and select workforce roles to deploy during the planning period – note, in this case study, this is a combination of two decisions made simultaneously |
| Team size and composition | Determine the number of staff members in each role that have to be available any day in the planning period in order to meet demand |
| Rostering & Allocation & Scheduling | For a given week in the planning period, decide which days each available staff member conducts visits, assign each staff member a set of patient visits and decide on which day of the week each visit has to be conducted – note, in this case study, this is a combination of three decisions made simultaneously |

### 4.2 Synthetic data generator

Based on the scheme represented in Figure 1, we need a dataset representing the health economy of interest from which instances for the mathematical programming formulations and heuristic algorithms described in the next section can be extracted. We identified the following two main groups of features that the dataset should contain to enable analysis of decision problems consistently throughout the three planning levels:

- Demand data. This is a list of patients with their geographical location and, for each patient, the required activities and frequencies for each week in the reference planning period (20 weeks in our analyses). The set of activities required weekly by a patient and their frequencies constitutes a “week profile”. Each patient can potentially switch each week from one profile to another profile (including a “profile zero” requiring no activities, i.e. patient not receiving any service).
- Service data. These include: a list of activities offered by the home health care organisation along with an associated service time; a list of potential workforce roles with salary rates; the subset of activities that a member of staff in each role may conduct based on their seniority level or other criteria; packing of activities into potential “visit types”, with corresponding visit times.

We designed the dataset structure to closely match the type of data usually available as (or easily obtainable from) routinely collected HHC patient records (Figure 4). However, especially for the analysis of strategic or tactical decisions and their effects on operational decisions, generating realistic but synthetic data with characteristics matching those of a real-world health economy of interest could be very useful. This approach would make exploration and comparison of different scenarios easier and support reproducible analyses accounting for stochastic variations in parameter values.

**Figure 4.**
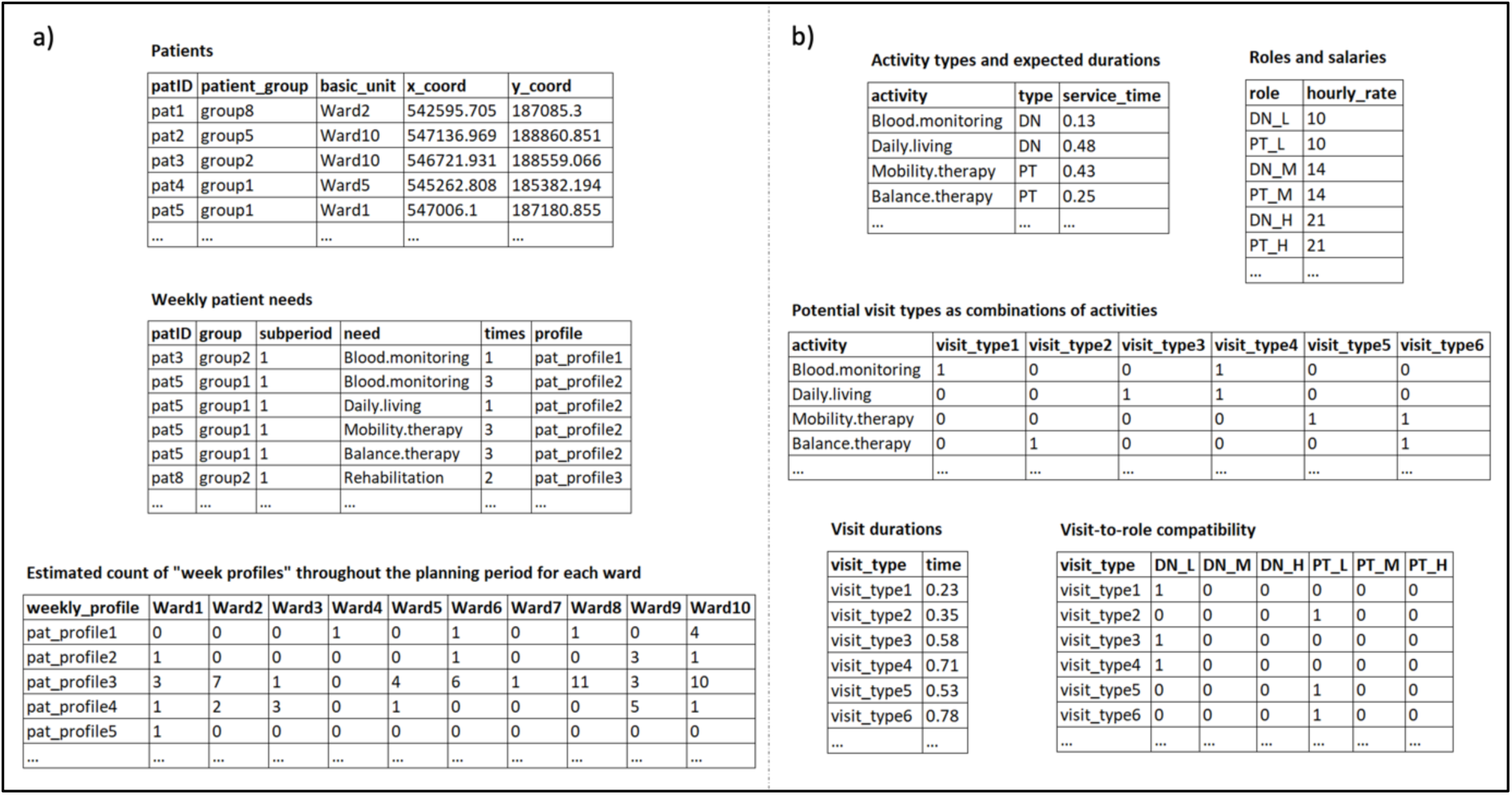
Example of structured synthetic dataset formed of demand features (a) and service features (b).

We developed an algorithm for the generation of synthetic data based on a set of input features for the health economy of interest. The algorithm, developed in R and available for download (Grieco et al. 2025), generates a dataset of the structure described above based on the following input information:

- A territory of interest, in the form of a set of wards with geographic coordinates defining their boundaries and demographic data (e.g. 65+ population). This is used to sample home locations of the synthetic patients. The synthetic data generator samples the total number of patients over the planning period from a discrete probability distribution (we assumed a Poisson distribution in this case study), therefore parameters for that distribution (i.e. expected number of patients in our case) have to be provided as input as well.
- Patient features. Initial probabilities of patients belonging to weekly profiles as well as transition probabilities between weekly profiles are used by the synthetic data generator to sample the evolution of weekly required activities and frequencies of activities for each synthetic patient. Transition probabilities can also be stratified into groups (for instance, based on referral reasons) in order to define synthetic patients based on different patterns of evolving need.
- Service features. These include the list of available activities and associated service times, the list of potential staff roles with corresponding salary rates, an overhead to account for activities not directly related to the performing of DN/PT tasks (e.g. establishing and developing a rapport with the patient) when computing visit times, and specification of which roles are allowed to conduct which activities.

### 4.3 Formulations

To quantitatively analyse the decisions in the hierarchy of interest, we defined one or more methods enabling computation of an optimal or heuristic solution for each step in the sequence and we implemented these methods into a configurable tool, available for download (Grieco et al. 2025), using R programming language. The tool can be used as it is to reproduce the analyses described in this paper, but its modular structure also allows for amendments of the decision framework and of the solution algorithms.

In this section, we provide brief descriptions of the formulations and heuristic procedures we developed for the case study presented in this paper (see Supplementary File 2 for more formal descriptions). In Table 5, for each decision considered, we list the upstream decisions (when applicable), the levers, the parameters derived from the underlying data and the output.

**Table 5.** Upstream decisions, levers, parameters and outputs for each decision considered in our analysis.

| Decision ($d$) | Upstream decisions ($U^d$) | Levers ($L^d$) | Parameters (retrieved from the underlying dataset $H$) | Output ($\bar{x}^d$) |
| --- | --- | --- | --- | --- |
| Districting | N/A | • Number of districts required<br>• Weight to be used for each component of the objective function<br>• Which pairs of basic units can/cannot belong to the same district (compatibility)<br>• Which pairs of basic units share borders (contiguity)<br>• Maximum distance allowed between any pair of basic units belonging to the same district<br>• Maximum allowed percentage deviation of workload in each district from average workload | • List of basic units<br>• Distance between centroids of each pair of basic units<br>• List of activities to be delivered over the planning horizon<br>• Activity durations<br>• Number of times each activity needs to be delivered in each basic unit over the planning horizon | • Set of basic units forming each district |
| Workforce roles & HHC packages | • Set of basic units forming each district (Districting) | • District(s) of interest | • List of potential staff roles<br>• List of potential visit types as combinations of activities<br>• List of weekly patient profiles<br>• Hourly salary cost for each potential role<br>• Duration of each potential visit type<br>• Set of weekly activities and frequencies associated with each weekly patient profile<br>• Number of times each weekly patient profile appears during the planning horizon<br>• List of potential visit types that can be conducted by each role | • Roles available for home visits<br>• Weekly frequency of a visit type being conducted by a given staff role to the benefit of a patient of a given weekly profile (Workforce roles & HHC packages) |
| Team size and composition | • Set of basic units forming each district (Districting)<br>• Roles available for home visits (Workforce roles & HHC packages)<br>• Weekly frequency of a visit type being conducted by a given staff role to the benefit of a patient of a given weekly profile (Workforce roles & HHC packages) | • District(s) of interest<br>• Minimum number viable of staff required for each role<br>• Assumed daily travel time for any staff member, for each role<br>• Proportion of daily scenario demand to cover for each visit type | • Maximum daily working hours for each role<br>• Fixed daily salary cost associated with the deployment of a staff member, for each role<br>• List of visit types that can be conducted by each role<br>• Number of visits of each type to be delivered each day in the planning horizon<br>• Visit durations | • Number of staff to be available daily, for each role<br>• Overall time spent on visits, for each role |
| Rostering & Allocation & Scheduling | • Set of basic units forming each district (Districting)<br>• Roles available for home visits (Workforce roles & HHC packages)<br>• Number of staff to be available daily, for each role (Team size and composition) | • District(s) of interest<br>• Weight to be used for each component of the objective function<br>• Maximum number of unique staff-patient pairs allowed over the planning period<br>• Minimum proportion of visits to be conducted over the planning period<br>• Monetary budget limit<br>• Average travel speed assumed for staff movement between patients | • List of patients requiring visits during the planning period<br>• Visit type and frequencies required by each patient over the planning period<br>• Distance between patient locations<br>• Visit durations<br>• List of visits that can be conducted by each staff member<br>• Fixed daily salary cost associated with the deployment of each staff member<br>• Maximum daily working hours for each staff member | • Days of the planning period in which each staff member is deployed for at least one visit<br>• Day of the planning period in which each visit is conducted and by which staff member<br>• Set of patients that each staff member visits at least once over the planning period |

#### 4.3.1 Districting

The reference territory is divided into a pre-defined number of districts (or clusters – 2 in our case study). In the districting integer programming formulation, adapted from Dugošija et al. 2020 and reported in Supplementary File 2, we consider the set of basic units (or wards) to be used as building blocks for a specified number of districts (or clusters), their pairwise distances (centroid to centroid), whether or not they share borders with each other (pairwise contiguity) and whether or not two basic units can belong to the same district for any reason (pairwise compatibility).

The expected workload in a basic unit is given by multiplying the estimated number of times a given activity will be requested in that basic unit over the planning period (20 weeks in our case study) by the expected duration of an activity of that type. Note, demand is considered at the level of activities and not at the level of whole visits, which are defined at a lower step in the hierarchy, and it excludes travel time.

In our case study, we optimised the balance of the expected workload required across the resulting districts. We measure the workload in terms of expected total activity time required to satisfy the annual demand. We define the deviation between each district’s total demand (in terms of annual activity time) and the average demand across districts (total demand divided by the envisaged number of districts). We minimise the maximum of those deviations to achieve a balance in the demand across the districts.

#### 4.3.2 Workforce roles & Home health care packages

This decision consists of selecting a set of workforce roles to deploy in a district over a given planning period (20 weeks in our case study), while combining the required activities into visit types. We defined a novel integer linear programming formulation for the simultaneous solution of these two decision problems (cf. Supplementary File 2). The formulation works at the level of “weekly patient profiles”, i.e. defining the set and composition of visits to be delivered to a given patient based on the set of activities the patient needs in a given week (in general, a single patient can “move” between different weekly profiles week by week).

Workforce roles and visit types are selected by determining the number of times each potential visit type should be conducted by each potential role to the benefit of patients of a given weekly profile over the planning period.

Staff roles are assigned to visit types at the same time as visit definition, based on visit-to-role constraints provided as input.

We solve the problem by minimising the total number of visits required to satisfy the demand over the planning period.

#### 4.3.3 Team size and composition

In our case study, we use a heuristic procedure to determine a daily team size to be available for home visits in a given district (cf. Supplementary File 2). In this simple decision-making algorithm, the team mix is determined by randomly assigning each required visit to one of the least expensive roles with appropriate seniority level for that visit type. To account for variability in the daily numbers of required visits, we sample daily demand scenarios for each week in the planning period. Then, for each visit type, we assign roles to visits in order to cover a given percentile (100% in our analyses) of the distribution of the daily number of visits.

#### 4.3.4 Rostering & Allocation & Scheduling

Our Rostering & Allocation & Scheduling formulation (cf. Supplementary File 2) allocates staff members to visits over the short term (planning period of 7 days in our case study) based on their seniority level, with a time resolution of 1 day (i.e. each staff member is assigned a set of visits each day, but without an hourly scheduling of visits). The demand is specified over a week (7 days) by the set of visits that each patient in a district should receive during that week.

In our analyses, we considered optimisation of the weighted sum of two components:

- Total weekly salary costs, formed of a daily fixed cost per salaried staff member, for the days that a staff member is deployed in at least one visit;
- Number of unique staff-patient pairs over the 7 days, which is used as a proxy for continuity of care.

### 4.4 Case study analysis

We conducted our analysis based on the diagram of Figure 3. The R code was set up to first execute the Districting decision, in particular clustering the administrative wards into two districts. For the sake of simplicity, we then continued the analysis focusing on one of the two districts only, as the characteristics of the geographic area and demand were homogeneous across all the wards and the modelling question did not require comparisons between different districts. For the chosen district (District 1), we ran the Workforce roles & HHC packages decision to select which roles to deploy and which visit types to consider based on the demand in the planning period of interest. We then defined the daily Team size and composition using our heuristic algorithm. Finally, we solved the Rostering & Allocation & Scheduling problem for a selected week in the planning period and summarised the results. We separately considered two possible objectives for the latter problem: minimisation of monetary costs and maximisation of continuity of care.

The above sequence of problems was analysed in two different scenarios:

- Scenario 1 – absence of the Hybrid Assistant in the set of possible roles;
- Scenario 2 – presence of the Hybrid Assistant in the set of possible roles.

The two scenarios were defined at the point of synthetic data generation, therefore two different datasets were produced, with and without the Hybrid Assistant, respectively. For a better comparison of the two scenarios, we duplicated all the demand-related data instead of re-generating them from scratch.

#### 4.4.1 Model parameterisation

We determined the input features for the synthetic data algorithm based on patient record data regarding our reference borough of London. By trimming the data to the period 01/01/2018 to 31/12/2018 and excluding records with missing key information, we obtained a dataset of 201,932 complete records covering 6,544 patients. Each record contained data including: referral labels, date/time of visit, staff conducting the visit and their role, activities carried out during the visit. To determine patient “week profiles” and needs, we classified patients into groups based on their sets of referral reasons (i.e. reasons for receiving care) during the planning period of interest. By discarding groups formed of fewer than 10 patients with the same set of referral reasons, we obtained 8 patient groups. For each patient group, we identified the set of relevant activities and computed the frequencies of switching between all the possible combinations of weekly activities (also considering the number of times an activity was delivered to the patient) which we used as proxies for the probabilities of a patient switching from one profile to another each week.

For the analyses described in this paper, we simplified the relevant features to create a smaller-scale synthetic dataset covering a planning period of 20 weeks and an expected number of 500 patients receiving visits at any time during the planning period. The generated data are available for download as R objects (Grieco et al. 2025).

#### 4.4.2 Analysis results

We ran the optimisation/heuristic routines described above according to the sequence specified in Figure 3 for Scenario 1 and Scenario 2 and compared the two scenarios using the performance metrics listed in Table 6.

**Table 6.** Performance metrics used for the assessment of the two scenarios of interest in our case study.

| Decision problem | Performance metric | Description |
| --- | --- | --- |
| Workforce roles & HHC packages | Total visits in the planning horizon | Number of visits that should be conducted during the whole planning period to satisfy the expected demand |
| Rostering & Allocation & Scheduling | Staff deployed weekly | Number of staff members that conduct at least one visit during the week of interest, overall and by role |
|  | Unique staff-patient pairs | Number of staff-patient pairs that meet at least once for a visit during the week of interest |
|  | Staff-per-patient ratio | Number of staff members visiting the same patient during the week of interest, averaged across all patients needing a visit on that week |
|  | Monetary costs | Sum of the daily salaries of all staff members deployed during the week of interest, each multiplied by the number of days in which the staff member conducts at least one visit |

The resulting districts obtained for both scenarios are shown in Figure 5. In our assessment, we mainly focused on the outputs for the decision problems “Workforce roles & HHC packages” and “Rostering & Allocation & Scheduling”.

**Figure 5.**
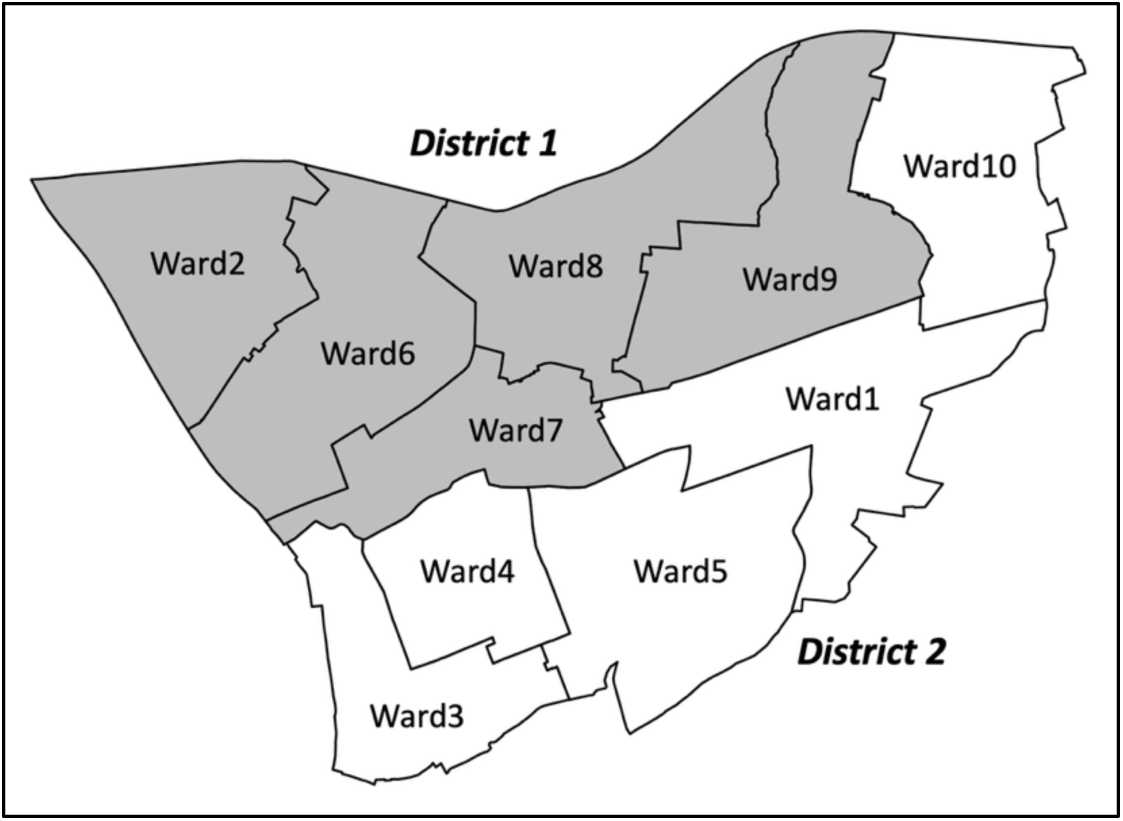
District 1 (grey) and District 2 (white) obtained as optimal solution of the Districting formulation balancing the estimated workload between the two resulting districts.

Analysis results are summarised in Figure 6 (optimisation of continuity of care at operational level) and Figure 7 (optimisation of monetary costs at operational level) for District 1.

**Figure 6.**
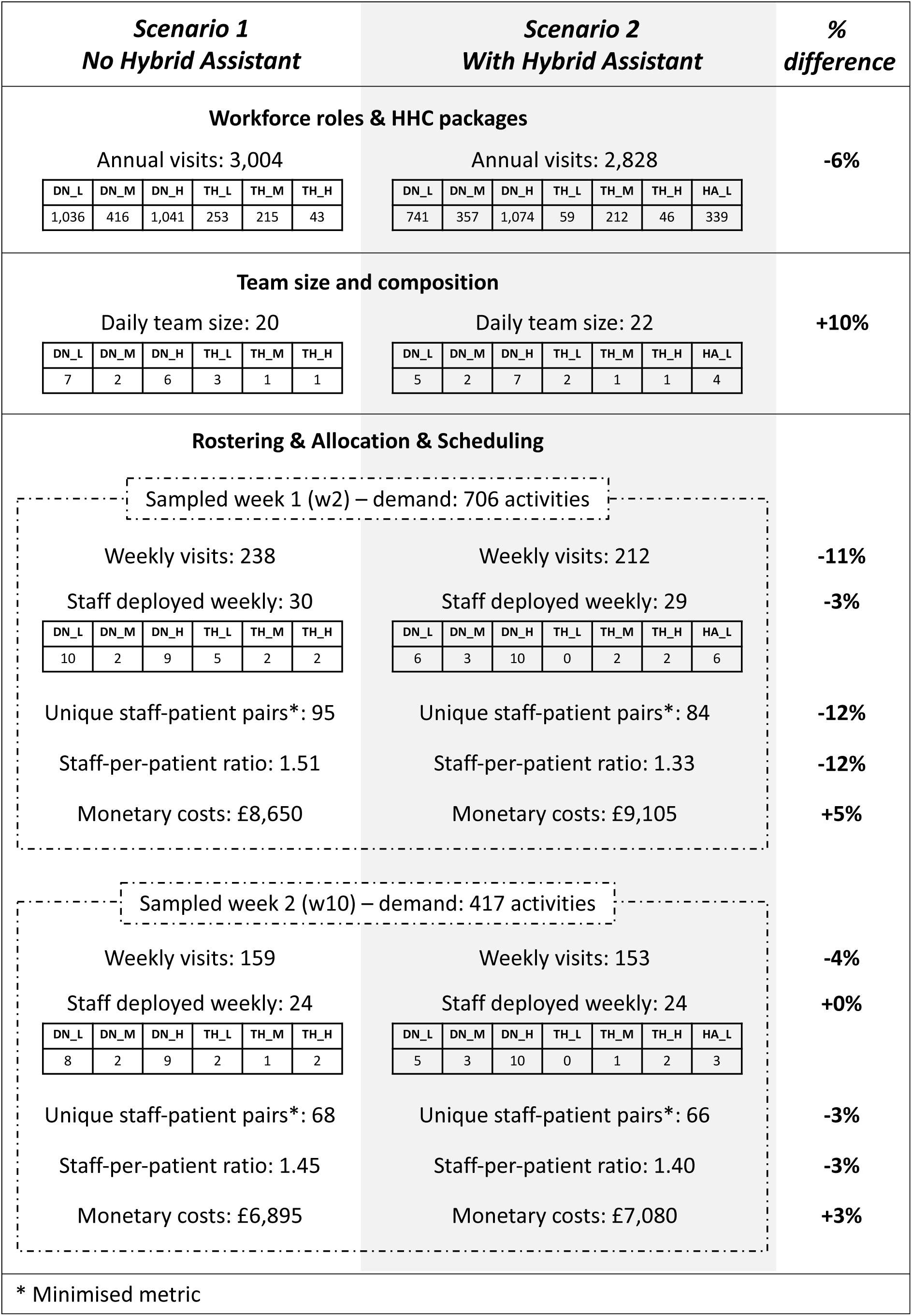
Results with optimisation of continuity of care for Rostering & Allocation & Scheduling. The sampled weeks correspond to the second (w2) and tenth (w10) weeks in the synthetic dataset.

**Figure 7.**
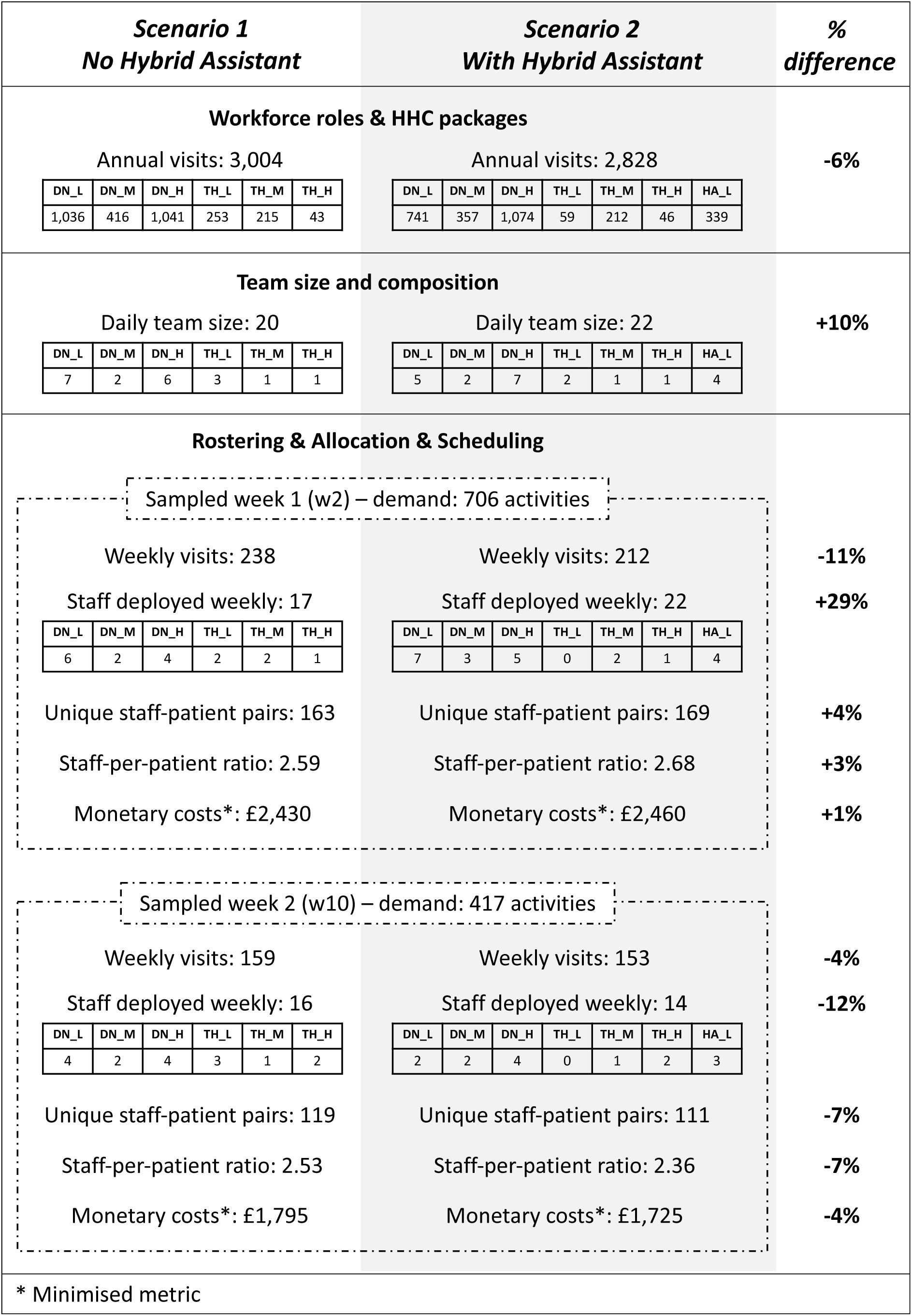
Results with optimisation of monetary costs for Rostering & Allocation & Scheduling. The sampled weeks correspond to the second (w2) and tenth (w10) weeks in the synthetic dataset.

Introduction of the Hybrid Assistant role would allow activities to be packed into visits in a more efficient way, with a resulting 6% decrease in the expected number of visits to be conducted to cover the same demand across the 20-week planning period.

We randomly sampled two weeks in the planning period to observe the optimal solutions obtained with the Rostering & Allocation & Scheduling formulation in the two scenarios, using a sub-optimal number of available staff members as obtained from application of the Team size and composition heuristic algorithm.

If the decision maker uses continuity-of-care criteria to optimise operational decisions (Figure 6), then the introduction of the Hybrid Assistant role results in a decrease in the number of weekly visits that are necessary to cover the whole demand (in line with the results from the Workforce roles & HHC packages problem), together with an improvement in the performance metrics measuring the level of continuity of care, namely the number of unique staff-patient pairs and the staff-per-patient ratio. These differences are accentuated for the week characterised by a higher number of weekly visits. In both cases, the total number of staff members deployed at least once during the week remains roughly the same in the two scenarios, but the total corresponding monetary costs are slightly higher with the new role.

If the decision maker aims instead to minimise monetary costs at an operational level (Figure 7), the presence of the Hybrid Assistant role has different effects at operational level in the two weeks analysed: it leads to a slight decrease in monetary costs in the week characterised by a lower number of visits, but a slight increase in monetary costs in the week characterised by a higher number of visits. In the latter case, the packing of activities into visit types conducted at strategic level might have hindered a less expensive allocation of staff to activities (e.g. the one obtained in the absence of the new role) at operational level – note, we assume that visit types are not modifiable once they have been decided in optimising Workforce roles & HHC packages.

## 5. Discussion

This study addresses a key gap in the HHC modelling literature by introducing a modelling approach for the analysis of decisions across multiple planning levels. Recent publications (Chabouh et al. 2023, Grieco et al. 2021), as well as our literature review update (section 2), identified a growing but very limited body of work on the modelling of HHC services at multiple planning levels, with existing studies focusing primarily on the development of mathematical formulations and solution algorithms for specific problems rather than creating reusable, generalisable tools. Our approach explicitly exploits the hierarchical nature of HHC decision-making while maintaining flexibility for application across diverse health care contexts.

A key contribution of this work lies in the modular design of the framework, which supports both reusability and adaptability. By structuring decisions as discrete but interlinked modules, the framework facilitates systematic analysis of how the decision variables, levers, outputs from upstream decisions, and health economy characteristics interact. This process could also be a useful exercise for the modeller and the stakeholders to identify and appropriately treat the controllable and non-controllable factors intervening in the decision process.

To enable practical adoption of our approach, we implemented it in R, providing a configurable modelling environment for the analysis of decision hierarchies. The code was structured to allow straightforward amendments, thereby encouraging adaptation by other modellers. We further developed a synthetic data generator to instantiate decision problems with consistent representations of health care system characteristics throughout the decision hierarchy of interest. Our synthetic data generator also contributes to addressing the gap identified by Chabouh et al. 2023 about the lack of benchmark instances for comparison of different methodologies, not necessarily focusing on the study of decision making at different planning levels but also regarding assessment of the multitude of scheduling/routing algorithms that have been developed – our tool can be easily adapted to generate data for such algorithms.

Despite these contributions, there are some limitations and areas for future work. The R tools we developed are prototypes and may lack the robustness required for immediate deployment in large-scale, real-world HHC systems. The case study was deliberately simplified to illustrate the feasibility of the approach. However, scaling to realistic problem sizes will require the development of tailored solution approaches (e.g. heuristic algorithms). Moreover, while the approach was designed with generalisability in mind, its development into software was informed primarily by a collaboration with a London-based HHC organisation, and validation in other health economies (for instance, non-UK systems) would be beneficial.

Therefore, future directions include testing the adaptability of our approach in different HHC contexts and for diverse combinations/hierarchies of decisions. Moreover, further development of our analysis tools into standalone software packages would contribute to wider adoption of the approach. At the same time, populating repositories of HHC decision models, including both formulations and solution algorithms, could create a shared community resource, paving the way for the development of integrated software environments capable of facilitating the specification, interlinking, and analysis of complex decision hierarchies in HHC.

## Data Availability

All data produced are available online at https://doi.org/10.5281/zenodo.18018852

