## Supplementary File 2 for "A modelling approach for the analysis of decisions in Home Health Care at multiple planning levels"

### Formulations

#### 1 Districting

The reference territory is divided into a pre-defined number of districts. In the districting integer programming formulation, adapted from Dugošija et al. 2020 (`https://doi.org/10.1007/s10479-020-03559-y`), we consider the set of basic units to be used as building blocks for a specified number of districts, their pairwise distances (centroid to centroid), whether or not they share borders with each other (pairwise contiguity) and whether or not two basic units can belong to the same district for any reason (pairwise compatibility).

##### 1.1 Sets

- $D$ = set of districts
- $U$ = set of basic units
- $H$ = set of activity types

##### 1.2 Parameters

- $w_c$ = weight assigned to compactness component of objective function
- $w_b$ = weight assigned to balance component of objective function
- $\lambda_{u,h}$ = estimated annual number of times activities of type $h \in H$ have to be conducted in basic unit $u \in U$
- $t_h$ = average duration of an activity of type $h \in H$
- $d_{u,v}$ = distance (in km) between basic units $u, v \in U$
- $d_{max}$ = maximum distance (in km) allowed between any pair of basic units in the same district
- $\tau$ = maximum allowed percentage deviation of workload in each district from average workload
- $\phi_{u,v}$ = compatibility index between basic units $u, v \in U$ (0 = incompatible, 1 = compatible)
- $\psi_{u,v}$ = basic unit contiguity: binary indicator if basic units $u, v \in U$ are contiguous

#### 1.3 Decision Variables

- $x_{u,j} \in \{0, 1\} = 1$ if basic unit $u \in U$ is assigned to district $j \in D$, 0 otherwise
- $d_{max} \geq 0 =$ auxiliary variables for compactness component of objective function
- $g_{max} \geq 0 =$ auxiliary variables for balance component of objective function
- $s_{u,j} \in \{0, 1\} =$ auxiliary variables for flow constraints
- $\alpha_{u,j} \geq 0 =$ auxiliary variables for flow constraints (integer)
- $\beta_{u,v,j} \geq 0 =$ auxiliary variables for flow constraints (integer)
- $\gamma_{u,j} \geq 0 =$ auxiliary variables for flow constraints (integer)
- $\delta_j \geq 0 =$ auxiliary variables for flow constraints (integer)
- $p_{u,v,j} \in \{0, 1\} =$ auxiliary variables for linearisation of flow constraints
- $r_{u,v,j} \in \{0, 1\} =$ auxiliary variables for linearisation of flow constraints

#### 1.4 Expressions of decision variables

- $W_j =$ workload in district $j \in D$: $W_j = \sum_{u \in U} \sum_{h \in H} \lambda_{u,h} \cdot t_h \cdot x_{u,j}$
- $\bar{W} =$ average workload across districts: $\bar{W} = \frac{1}{|D|} \sum_{u \in U} \sum_{h \in H} \lambda_{u,h} \cdot t_h$

#### 1.5 Objective Function

**Minimise weighted combination of compactness and balance:**

$$\min \quad w_c \cdot d_{max} + w_b \cdot g_{max} \tag{1}$$

Optimises district formation by balancing compactness (minimising maximum distance within districts) and workload balance (minimising maximum deviation from average workload).

#### 1.6 Constraints

**Balance auxiliary variable constraints:**

$$g_{max} \geq \frac{W_j - \bar{W}}{\bar{W}} \quad \forall j \in D \tag{2}$$

$$g_{max} \geq \frac{\bar{W} - W_j}{\bar{W}} \quad \forall j \in D \tag{3}$$

Link the balance auxiliary variable to capture the maximum relative deviation of any district's workload from the average workload.

**Compactness auxiliary variable constraints:**

$$d_{max} \geq d_{u,v} \cdot (x_{u,j} + x_{v,j} - 1) \quad \forall u, v \in U, \forall j \in D \tag{4}$$

Link the compactness auxiliary variable to capture the maximum distance between any pair of basic units within the same district.

**Assignment constraints:**

$$\sum_{j \in D} x_{u,j} = 1 \quad \forall u \in U \tag{5}$$

Ensure that each basic unit is assigned to exactly one district.

**Compatibility constraints:**

$$(1 - \phi_{u,v}) \cdot (x_{u,j} + x_{v,j}) \leq 1 \quad \forall u, v \in U, \forall j \in D \tag{6}$$

Prevent incompatible basic units from being assigned to the same district.

**Distance constraints:**

$$d_{u,v} \cdot (x_{u,j} + x_{v,j} - 1) \leq d_{max} \quad \forall u, v \in U, \forall j \in D \tag{7}$$

Ensure that basic units assigned to the same district are not too far from each other when workload balance is optimised.

**Workload balance constraints:**

$$(1 - \tau) \cdot \bar{W} \leq W_j \quad \forall j \in D \tag{8}$$

$$W_j \leq (1 + \tau) \cdot \bar{W} \quad \forall j \in D \tag{9}$$

Ensure that the workload in each district does not deviate too much from the average workload when district compactness is optimised.

**Flow constraints for connectivity:**

$$\sum_{u \in U} s_{u,j} = 1 \quad \forall j \in D \tag{10}$$

$$s_{u,j} \leq x_{u,j} \quad \forall u \in U, \forall j \in D \tag{11}$$

$$\alpha_{u,j} \leq |U| \cdot x_{u,j} \quad \forall u \in U, \forall j \in D \tag{12}$$

$$\gamma_{u,j} \leq x_{u,j} \quad \forall u \in U, \forall j \in D \tag{13}$$

$$\sum_{u \in U, j \in D} \alpha_{u,j} = |U| \tag{14}$$

$$\sum_{u \in U} \gamma_{u,j} = \delta_j \quad \forall j \in D \tag{15}$$

Establishe flow relationships between auxiliary variables to ensure district connectivity and proper flow distribution.

**Flow balance constraints:**

$$\alpha_{l,j} + \sum_{u \in U} \beta_{u,l,j} \cdot \psi_{u,l} = \gamma_{l,j} + \sum_{v \in U} \beta_{l,v,j} \cdot \psi_{l,v} \quad \forall l \in U, \forall j \in D \tag{16}$$

Ensure flow conservation at each basic unit within each district, maintaining connectivity through contiguous units.

**Linearisation constraints:**

$$\alpha_{u,j} \leq |U| \cdot r_{u,u,j} \quad \forall u \in U, \forall j \in D \tag{17}$$

$$x_{u,j} + s_{u,j} - 1 \leq 2 \cdot r_{u,u,j} \quad \forall u \in U, \forall j \in D \tag{18}$$

$$2 \cdot r_{u,u,j} \leq x_{u,j} + s_{u,j} \quad \forall u \in U, \forall j \in D \tag{19}$$

$$\beta_{u,v,j} \leq |U| \cdot p_{u,v,j} \cdot \psi_{u,v} \quad \forall u, v \in U, \forall j \in D \tag{20}$$

$$x_{u,j} + x_{v,j} - 1 \leq 2 \cdot p_{u,v,j} \quad \forall u, v \in U, \forall j \in D \tag{21}$$

$$2 \cdot p_{u,v,j} \leq x_{u,j} + x_{v,j} \quad \forall u, v \in U, \forall j \in D \tag{22}$$

Linearise the relationships between flow variables and assignment variables using binary auxiliary variables.

### 2 Workforce roles & HHC packages

This decision consists of selecting a set of workforce roles to deploy in a district over a given planning period, while combining the required activities into visit types. This integer-linear programming formulation works at the level of "weekly patient profiles", i.e. defining the set and composition of visits to be delivered to a given patient based on the set of activities the patient needs in a given week (in general, a single patient can "move" between different weekly profiles week by week). Staff roles are assigned to visits at the same time as visit definition, based on "role-to-visit" constraints provided as input. The problem can be solved by minimising either the total number of visits or the total staff costs (based on hourly salaries) required to satisfy the demand over the planning period.

#### 2.1 Sets

- $P$ = set of weekly patient profiles
- $V$ = set of visit types
- $A$ = set of activities
- $R$ = set of roles

#### 2.2 Parameters

- $h_r$ = hourly rate (£) of role $r \in R$
- $d_v$ = duration (hours) of visit type $v \in V$
- $c_{a,v}$ = number of times activity $a \in A$ is performed in visit type $v \in V$
- $\delta_{p,a}$ = number of times a patient of profile $p \in P$ needs activity $a \in A$ during a week
- $\rho_{r,v}$ = role-to-visit compatibility: binary indicator denoting whether a staff member in role $r \in R$ can conduct a visit of type $v \in V$
- $n_p$ = number of patients belonging to the weekly profile $p \in P$
- $M$ = large constant = $\sum_{p \in P} \sum_{a \in A} \delta_{p,a}$

#### 2.3 Decision Variables

- $x_{v,p,r} \geq 0$ = number of times a visit of type $v \in V$ is conducted by a staff member in role $r \in R$ for the benefit of a patient of profile $p \in P$ over a week (integer)

#### 2.4 Objective Function

**Minimise total number of visits:**

$$\min \quad \sum_{v \in V} \sum_{p \in P} \sum_{r \in R} n_p \cdot x_{v,p,r} \tag{23}$$

Minimises the total number of healthcare visits across all patients to achieve operational efficiency.

**Alternative objective (minimise total treatment cost):**

$$\min \quad \sum_{v \in V} \sum_{p \in P} \sum_{r \in R} n_p \cdot h_r \cdot d_v \cdot x_{v,p,r} \tag{24}$$

Minimises the total monetary cost of healthcare delivery based on staff hourly rates and visit durations.

#### 2.5 Constraints

**Demand satisfaction constraints:**

$$\sum_{r \in R} \sum_{v \in V} c_{a,v} \cdot x_{v,p,r} = \delta_{p,a} \quad \forall p \in P, \forall a \in A \tag{25}$$

Ensure that each patient profile receives exactly the required number of each healthcare activity per week.

**Role-visit compatibility constraints:**

$$x_{v,p,r} \leq \rho_{r,v} \cdot M \quad \forall v \in V, \forall p \in P, \forall r \in R \tag{26}$$

Prevent assignment of visit types to staff roles that are not qualified or authorised to perform them.

**Non-negativity and integrality:**

$$x_{v,p,r} \geq 0 \text{ and integer} \quad \forall v \in V, \forall p \in P, \forall r \in R \tag{27}$$

### 3 Team size and composition

We defined a heuristic procedure to determine a sub-optimal daily team size to be available for home visits in a given district. In this simple decision-making algorithm, the team mix is determined by randomly assigning each required visit to one of the cheapest roles with appropriate skills for that visit type. To account for variability in the daily numbers of required visits, we consider a planning period formed of a given number of daily demand scenarios. Then, for each visit type, we assign roles to visits in order to cover a given percentile of the distribution of the number of daily visits across the scenarios.

#### 3.1 Parameters

- Set of roles $R$ (each denoted with index $r$)
- Set of visit types $V$ (each denoted with index $v$)
- Set of roles that can conduct visits of type $v$: $R_v \subseteq R$
- Set of daily demand scenarios (number of visits) for visit type $v$: $D_v \in \mathbb{N}$
- Percentile of demand to cover over the whole planning period (same for each visit type): $p \in \mathbb{R}, \geq 0$
- Maximum daily working hours (including travel time) per salaried staff member in role $r$: $\Sigma_r \in \mathbb{R}, \geq 0$
- Estimated daily travel time per salaried staff member in role $r$: $t_r \in \mathbb{R}, \geq 0$
- Service time (in hours) for any visit of type $v$: $\delta_v \in \mathbb{R}, \geq 0$
- Daily deployment cost (in £) for each salaried staff member in role $r$: $s_r \in \mathbb{R}, \geq 0$

#### 3.2 Decision variables

- Number of salaried staff deployed daily per role: $n_r \in \mathbb{N}$
- Daily workload (excluding estimated travel time) of all staff members deployed in each role: $W_r \in \mathbb{R}, \geq 0$

#### 3.3 Algorithm steps

*Initialisation*

- Set daily workload for each role to zero:

$$W_r := 0, \quad \forall r \in R$$

*Phase 1*

For each visit type $v \in V$:

- Determine the reference daily demand $d_v$ corresponding to the percentile $p$ of the daily demand scenarios $D_v$
- Assign the cheapest role to visit type $v$ among the roles that can conduct it:

$$r^* := \arg\min_r \left\{ \frac{s_r}{\Sigma_r} : r \in R_v \right\}$$

- Update the daily workload for the selected role:

$$W_{r^*} := W_{r^*} + d_v \cdot \delta_v$$

*Phase 2*

For each role $r \in R$:

- Set number of salaried staff members required daily:

$$n_r := \max \left\{ 0, \left\lceil \frac{W_r}{\Sigma_r - t_r} \right\rceil \right\}$$

### 4 Rostering & Allocation & Scheduling

This integer-linear programming formulation allocates visits to staff members over the short term (for instance, planning period of 7 days in our case study) based on their skills, with a time resolution of 1 day (i.e. each staff member is assigned a set of visits each day, but without an hourly scheduling of visits). The demand is specified over the planning period by the set of visits that each patient in a district should receive during that period. The formulation also includes constraints around the minimum and maximum elapsed time between visits to the same patient. The objective function considers the balance between to components: the total weekly salary costs, formed of a daily fixed cost per salaried staff member, for the days that a staff member is deployed in at least one visit; the number of unique patient-staff pairs over the planning period, which is used as a metric for continuity of care.
Note, while this formulation also decides in which day each staff member will need to be available (roster), it relies on an input specifying days in which each staff member could be possibly deployed. For instance, in our case study, we allow each staff member to be available for no more than five days across a seven-day planning period. Therefore, in our implementation, we have included a heuristic routine in the construction of the input to the this formulation that determines on which days each staff member could be selected: starting from the required team size and composition, we generate the corresponding minimum number of staff members required in each role and assuming that each staff member is available for no more than five days a week, and then we randomly assign staff members to days such that the team size and composition is constant throughout the week.

#### 4.1 Sets

- $P$ = set of patients
- $S$ = set of staff members
- $R$ = set of roles
- $T$ = set of time points identifying sub-periods (day 1, day 2, etc.)
- $V$ = set of visits to be conducted during the planning period

#### 4.2 Parameters

- $w_c$ = weight assigned to costs component of objective function
- $w_p$ = weight assigned to unique pairs component of objective function
- $A_{s,t}$ = binary indicator if staff member $s \in S$ is available during sub-period $t \in T$
- $\nu_{v,p}$ = binary indicator if visit $v \in V$ belongs to patient $p \in P$
- $M_p^{daily}$ = maximum number of daily visits allowed per patient $p \in P$
- $\kappa_{v,s}$ = binary indicator if staff member $s \in S$ can conduct visit $v \in V$

- $\tau_v$ = service time for visit $v \in V$
- $\tau^{travel}$ = average travel time between any pair of locations in the same district
- $W_s^{max}$ = maximum working time (including travel time) for staff member $s \in S$ in each sub-period
- $c_s$ = daily salary of staff member $s \in S$
- $\phi^{min}$ = minimum proportion of visits to be satisfied over the planning period
- $N^{max}$ = maximum number of unique staff-patient pairs allowed
- $B^{limit}$ = budget limit
- $\rho_{s,r}$ = binary indicator if staff member $s \in S$ belongs to role $r \in R$

#### 4.3 Decision Variables

- $x_{s,v,t} \in \{0, 1\} = 1$ if staff member $s \in S$ is allocated to visit $v \in V$ during sub-period $t \in T$, 0 otherwise
- $y_{s,p} \in \{0, 1\} = 1$ if staff member $s \in S$ visits patient $p \in P$ at least once during the planning period, 0 otherwise
- $z_{s,t} \in \{0, 1\} = 1$ if staff member $s \in S$ is allocated at least one visit during sub-period $t \in T$, 0 otherwise

#### 4.4 Expressions

- $W_{s,t}$ = daily workload of staff member $s \in S$ in sub-period $t \in T$:

$$W_{s,t} = \sum_{v \in V} (\tau_v + \tau^{travel}) \cdot x_{s,v,t} + \tau^{travel} \cdot z_{s,t}$$

- $W_s$ = overall workload of staff member $s \in S$: $W_s = \sum_{t \in T} W_{s,t}$
- $\Phi$ = proportion of visits conducted: $\Phi = \frac{\sum_{s \in S, v \in V, t \in T} x_{s,v,t}}{|V|}$
- $N$ = number of unique staff-patient pairs: $N = \sum_{s \in S, p \in P} y_{s,p}$
- $C$ = monetary costs: $C = \sum_{s \in S, t \in T} c_s \cdot z_{s,t}$

#### 4.5 Objective Function

**Minimise weighted combination of costs and unique staff-patient pairs:**

$$\min \quad w_c \cdot C + w_p \cdot N \tag{28}$$

Optimises home-based healthcare delivery by balancing monetary costs (minimising salary costs) and continuity of care (minimising unique staff-patient pairs).

#### 4.6 Constraints

**Staff-patient relationship constraints:**

$$y_{s,p} \geq \nu_{v,p} \cdot x_{s,v,t} \quad \forall s \in S, \forall p \in P, \forall v \in V, \forall t \in T \tag{29}$$

$$y_{s,p} \leq \sum_{v \in V, t \in T} \nu_{v,p} \cdot x_{s,v,t} \quad \forall s \in S, \forall p \in P \tag{30}$$

Lin staff-patient relationship variables to visit allocation variables, ensuring proper tracking of which staff members serve which patients.

**Staff deployment constraints:**

$$z_{s,t} \geq x_{s,v,t} \quad \forall s \in S, \forall t \in T, \forall v \in V \tag{31}$$

$$z_{s,t} \leq \sum_{v \in V} x_{s,v,t} \quad \forall s \in S, \forall t \in T \tag{32}$$

Link staff deployment variables to visit allocation variables, tracking when staff members are active in each sub-period.

**Visit uniqueness constraints:**

$$\sum_{s \in S, t \in T} x_{s,v,t} \leq 1 \quad \forall v \in V \tag{33}$$

Ensure that each visit is conducted at most once across all staff members and time periods.

**Staff availability constraints:**

$$z_{s,t} \leq A_{s,t} \quad \forall s \in S, \forall t \in T \tag{34}$$

Prevent staff deployment when they are not available for work.

**Staff skill compatibility constraints:**

$$\sum_{t \in T} x_{s,v,t} \leq \kappa_{v,s} \quad \forall v \in V, \forall s \in S \tag{35}$$

Ensure that staff members are only allocated to visits they are qualified to perform.

**Working time limit constraints:**

$$W_{s,t} \leq W_s^{max} \quad \forall s \in S, \forall t \in T \tag{36}$$

Enforce maximum working time limits for each staff member in each sub-period, including service and travel time.

**Daily visit balance constraints:**

$$\sum_{s \in S, v \in V} \nu_{v,p} \cdot x_{s,v,t} \leq M_p^{daily} \quad \forall p \in P, \forall t \in T \tag{37}$$

Keep a balanced distribution of visits to each patient across the planning period.

**Minimum visit coverage constraint:**

$$\Phi \geq \phi^{min} \tag{38}$$

Ensure that a minimum proportion of all required visits are successfully scheduled and allocated.
